# Post-surgery level of circulating DNA in stage III colon cancer patients: impact on the reliability of minimal residual disease detection

**DOI:** 10.1101/2025.04.04.25324955

**Authors:** Andrei Kudriavtsev, Saidi Daoud, Catalina Isabel Cofre Muñoz, Alexia Mirandola, Ekaterina Pisareva, Javier Gonzalo Ruiz, Marco Macagno, Nadia Saoudi Gonzalez, Evelyne Crapez, Marc Ychou, Ramon Salazar Soler, Elisabetta Fenocchio, Paula X. Fernandez Calotti, Thibault Mazard, Cristina Santos Vivas, Elena Elez, Federica Di Nicolantonio, Alain R. Thierry

**Author notes:** Corresponding author: Alain R. Thierry, IRCM, Montpellier Cancer Research Institute, INSERM U1194, Montpellier University, Institut régional du Cancer de Montpellier, Montpellier, F-34298, France. These authors contributed equally to this work and share first authorship (Andrei Kudriavtsev, Saidi Daoud).

## Abstract

**Purpose:** Minimal residual disease (MRD) assessment guided by circulating DNA (cir-nDNA) testing is strongly prognostic in operable colon cancer patients. However, further clinical research to result in fully changing the clinical management of colon cancer about the use of adjuvant therapy.

**Experimental Design:** In order to improve the analytical signal sensitivity of MRD detection we investigated the sources of cir-nDNA and its concentration variation in 74 stage III colon cancer patients before surgery and up to eight weeks after tumor resection.

**Results:** A majority of stage III colon cancer patients showed significant higher post-surgery cir-nDNA levels as compared to pre-surgery level (82.2% and 64.4% during the first month and second month period). We observed a strong association of two neutrophil extracellular traps (NETs) markers (Myeloperoxidase, MPO and neutrophil elastase, NE) during the two-months post-operative period.

**Conclusions:** Whereas the literature previously assumed that cir-nDNA concentration significantly decreased at most one-month post-surgery in the majority of stage III colon cancer patients, our data challenge this paradigm. NETs appear to constitute a confounding factor in assessing total cir-nDNA concentration since NETs production largely varies post-surgery among patients. Thus, data suggest that it would be misleading to define an optimal post-surgery blood collection time for MRD detection. At best, we estimate that given the high level of cir-nDNA content and large inter-individual variability, the best time range for blood collection could be between the fourth and the sixth week post-surgery. Second, the value of the variant allele frequency (VAF) that is so far a criteria to select mutant cir-nDNA, should be cautiously taken into consideration. We are providing various recommendations in that regards.

**Translational Relevance:** Our observation is of translational relevance with regard to the future practice of cancer medicine, specifically minimal residual disease (MRD) assessment guided by cir-nDNA testing, which shows considerable promise in that domain. Given high inter-individual variation, as well as the high levels of cir-nDNA concentration arising from neutrophil extracellular traps (NETs) formation, we conclude that no optimal post-surgery blood collection time for MRD detection can be determined for all patients, and the use of the mutation allele frequency (MAF) should be limited. Our data suggest that best practice would be to accurately test for cir-nDNA levels, to monitor for NETs-derived inflammation, and to use the absolute mutant DNA quantification instead of MAF in the post-operative period.

## Introduction

Conventional histopathological and radiological examinations show limitations in determining which patients have minimal residual disease (MRD) (1)(2). Molecular MRD testing can support more individualized decisions about patient management, particularly as to the probable efficacy of treatment, in the early detection of recurrence, and with regard to prognosis(3)(4). The standard care management of patients with stage III colon cancer, or of high-risk stage II patients, includes surgery followed by adjuvant chemotherapy (ACT) (1)(2). However, about 15% and 30% of stage II and stage III colon cancer patients relapse following ACT, whereas an estimated 80% and 50% are cured by surgery only, respectively (4). Consequently, it remains a major challenge to accurately identify those patients who ought to be administered ACT or a lower ACT dose regimen (1)(2).

MRD assessment would identify patients who could benefit from adjuvant therapy, while a failure to detect MRD would spare patients who have already undergone surgery from having to also undergo adjuvant therapy and the ensuing toxicities (1)(2)(3)(4). The detection of circulating DNA derived from malignant cells would appear to be a promising method of predicting relapse in many solid cancers (3)(4)(5)(6). Along with an increased exploitation of circulating DNA (cir-nDNA, (7)) in a theragnostic strategy (8), MRD detection via this newly-identified biological source has obvious advantages. Pioneering work in this field by Tie and colleagues (2),(9) confirmed the benefits of stratifying patients using cir-nDNA for detecting residual disease, particularly those with stage II and III colon cancer.

In patients with stage III colon cancer, it is clinical practice starting adjuvant chemotherapy between the second and the eighth weeks post-surgery. Great advances have been made in evaluating the performance of adjuvant therapy guided by MRD detection as determined by cir-nDNA analysis (2)(5)(9). However, all assays have false positive and false negative cases, and in a fraction of individuals cir-nDNA detection is not evaluable (3)(4), suggesting the need for further clinical research in this area. We hypothesize that novel insights could be derived from research that considers the individualization of data for each patient, and confounding factors with respect to the analytical signal sensitivity of MRD detection. To this end, we studied the variations and the origins of cir-nDNA up to eight weeks post-surgery in each stage III colon cancer patient of the THRuST clinical study. Given that we previously showed that increases in cir-nDNA are associated with neutrophil extracellular traps (NETs) formation in mCRC patients at diagnosis (10)(11), here we evaluated the impact of NETs formation on the variation of cir-nDNA amount within this post-surgery period.

## Materials and Methods

### Patients and cohort

In this work, we examined plasma from all stage III colon cancer patients included in the THRuST clinical study (015-FPO18), which was a prospective, multicenter, and blinded observational study. THRuST is an ERC Transcan European project (https://www.transcanfp7.eu/index.php/abstract/thrust.html). The inclusion criteria selected patients aged ≥18 years old with histologically confirmed stage III colorectal adenocarcinoma. The main exclusion criteria were: active viral infection (hepatitis, HPV, HIV), previous systemic or radiation therapy for colorectal cancer, and a history of another neoplastic disease. The study’s primary objective was to assess the clinical feasibility of dynamically detecting tumour progression by monitoring a molecular and personalized signature by means of a blood test. The study’s secondary objectives included: an examination of the performance of each qualitative and quantitative cir-nDNA parameter, the acquisition of descriptive knowledge about the clonal evolution of driver mutations under standard care, and the comparison of the data with conventional biomarkers and imaging. Patients were screened and included at the IRCC (Istituto di Candiolo-Fondazione del Piemonte per l’Oncologia, Candiolo, Italy), VHIO (Vall d’Hebron University Hospital, Vall d’Hebron Institute of Oncology, Barcelona, Spain), ICO (Catalan Institute of Oncology, L’Hospitalet de Llobregat, Barcelona, Spain) and ICM (Montpellier Cancer Institute, Montpellier, France).

In the THRuST protocol, individual molecular signatures were defined for each CRC patient, based on next-generation sequencing analyses performed on DNA from tumour tissue after resection. Thus, this individual molecular tag consisted of at least one actionable driver, one non-actionable driver and a passenger mutation. The IntPlex method was employed for patient follow-up, enabling simultaneous determination of five parameters: (i) cir-nDNA concentration, (ii) presence of a point mutation, (iii) mutant DNA concentration, (iv) mutant allele fractions of cir-nDNA, and (v) cir-nDNA fragmentation index. In addition, specific cir-nDNA methylation patterns were monitored post-surgery. Blood samples for this study were collected before and after surgery, and at each follow-up visit (every 3 months if possible). The collected data was analysed with reference to clinical observations, standard management care, conventional imaging methods, and conventional CRC biomarkers. In this ancillary study, we only examined the dynamics of cir-nDNA in the course of the THRuST clinical study.

The protocol was approved by Ethics Committee of Candiolo Cancer Institute FPO-IRCCS and then by all participating institutions. All patients gave their written informed consent before enrolment. The study was conducted in accordance with the Declaration of Helsinki.

The healthy individual control cohort was composed of samples from blood donors to the Etablissement Français du Sang (EFS, Montpellier, France). EFS blood samples are highly controlled and qualified. They are subjected to the same preanalytical conditions as for patients’ blood samples before plasma analysis.

### Blood samples collection

Each clinical center followed the same strict and precise guidelines(16) concerning blood collection, tube handling and storage(16). Each center has a specific certified and registered CC bank, which adheres to the THRuST clinical study protocol (015-FPO18). Ethylene diamine tetraacetic acid tubes (EDTA) plasma samples were stored at −80C° and were periodically sent to both the IRCM (3 mL plasma minimum) and the IRCCS (1 tube, 3 mL plasma minimum). In receiving these samples, the IRCM team used quality controls as defined by existing cir-nDNA preanalytic guidelines(17). Blood samples were collected between Avril 2019 and July 2023. In the course of their routine pre- and post-surgical surveillance, patients were submitted to a 10-point blood sampling plan: 1 pre-surgery, 1 post-surgery (15<d<40 days), and every 3 months thereafter, concomitant with the obtention of clinical and imaging data, all of which is being collected in a customized eCRF. Each patient will be followed up pro-actively for a duration of up to 24 months or until disease recurrence, whichever comes first.

### Blood samples collection

Each clinical center followed the same strict and precise guidelines(16) concerning blood collection, tube handling and storage(16). Each center has a specific certified and registered CC bank, which adheres to the THRuST clinical study protocol (015-FPO18). Ethylene diamine tetraacetic acid tubes (EDTA) plasma samples were stored at −80C° and were periodically sent to both the IRCM (3 mL plasma minimum) and the IRCCS (1 tube, 3 mL plasma minimum). In receiving these samples, the IRCM team used quality controls as defined by existing cir-nDNA preanalytic guidelines(17). Blood samples were collected between Avril 2019 and July 2023. In the course of their routine pre- and post-surgical surveillance, patients were submitted to a 10-point blood sampling plan: 1 pre-surgery, 1 post-surgery (15<d<40 days), and every 3 months thereafter, concomitant with the obtention of clinical and imaging data, all of which is being collected in a customized eCRF. Each patient will be followed up pro-actively for a duration of up to 24 months or until disease recurrence, whichever comes first.

### Plasma isolation and cir-nDNA extraction

EDTA tubes were centrifuged at 1200 g for 10 min at 4 °C, within 4 h of collection. Plasma samples were immediately stored at 80 °C and transferred on dry ice from the recruiting institutions to our laboratory. Plasma was stored for a number of months (4–12 months), and centrifuged at 16 000 g for 10 min at 4 °C. An aliquot of plasma was then used to perform an enzyme-linked immunosorbent assay. Cir-nDNA was extracted from 1 mL plasma (Maxwell_ RSC Instrument) using the cfDNA Plasma Kit (Promega Corporation, Madison, WI, USA) in an elution volume of 130 μL. For our quantification of cir-nDNA, we adhered strictly to the cir-nDNA preanalytical guidelines referred to above (plasma isolation, plasma storage at −80°C(16), (17)). DNA extracts were stored at −20 °C until use. In storage at −80 °C(18) for up to 3 years, no significant variation of cir-nDNA concentration was previously reported as determined by quantification by Q-PCR a WT sequence of *KRAS* (67 bp length) used in this study. In total, 357 serial plasma samples from 74 patients were analyzed.

### Quantification of cir-nDNA

Analysis of cir-nDNA was carried out using the multiplexed IntPlex® method which was specifically designed for quantifying cir-nDNA(19). Briefly, on a CFX96 instrument using the CFX manager software (Bio-Rad), Q-PCR amplifications were conducted in two replicates, with each reaction having a total volume of 25 µL. Each PCR reaction comprised of 12.5 µL of IQ Supermix Sybr Green (Bio-Rad), 2.5 µL of DNase-free water (Qiagen) or specific oligoblocker, 2.5 µL of forward and reverse primers (0.3 pmol/mL), and 5 µL of DNA extract. The thermal cycling process consisted of three repeated steps: a hot-start activation step at 95 °C for 3 minutes, followed by 40 denaturation–amplification cycles at 95 °C for 10 seconds, then at 60°C for 30 seconds. Melting curves were studied by gradually increasing the temperature from 60°C to 90°C, with a plate reading taken at every 0.2°C increment. With a genomic extract of the DiFi cell line at 1.8 ng/mL of DNA, standards curves were generated for each run, in order to maintain accuracy and consistency in the results. Each PCR run was carried out with template control and positive control for each primer set. Validations of Q-PCR amplification were controlled by melt curve differentiation. To quantify the total circulating nuclear DNA (cir-nDNA) concentration in both CC patients and HI, amplification of a 67 bp-length wild-type sequence of the KRAS gene was performed. The coefficient of variation was determined as 24% for the quantification of cir-nDNA, when considering variation due to the extraction procedure and analysis in the same plate(20). The accuracy of this study’s measurement of cir-nDNA concentration is supported by two assessments: (i) the total cir-nDNA concentration, obtained by targeting a KRAS sequence, was routinely controlled by quantifying a BRAF internal control sequence. In addition, this quality control enabled the detection and exclusion of samples which presented a loss of heterozygosity (LOH) or gene amplification, two phenomena which have been reported in CRC patients(21). Moreover, since KRAS amplification is an infrequent event in CRC (0.67%)(21), its level did not impact our observations or the values described here; and (ii) this method of quantifying cir-nDNA has undergone rigorous experimental and clinical validation, demonstrating unparalleled specificity and sensitivity, to the point of permitting the detection of a single DNA fragment molecule, as determined under Poisson Law distribution experiment. A healthy cir-nDNA median value (12.6 ng/mL) was determined using a control cohort of 22 healthy individuals (Suppl. Fig.1). To facilitate our observations, we arbitrarily defined a positivity threshold for cir-nDNA concentration, thus enabling us to rigorously distinguish pathological or abnormal values with control values by adding the standard deviation to the median value of the healthy individuals (12.6 ng/mL). Note, we quantified plasma DNA by targeting a nuclear DNA sequence.

**Figure 1:**
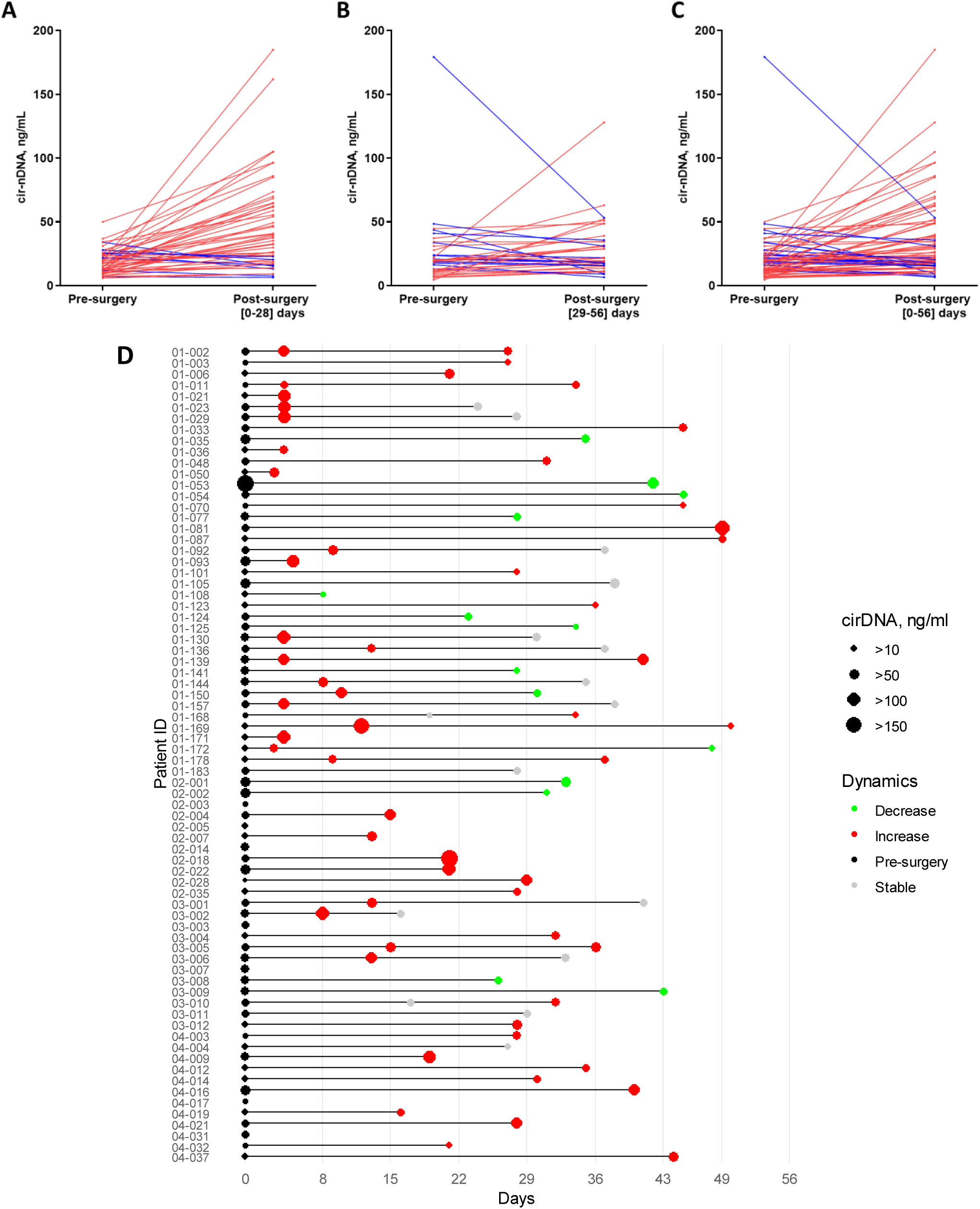
Comparative analysis of cir-nDNA concentrations in pre-surgery and post-surgery. The graphs representing the comparative analysis of pre-surgical versus post-surgical values of cir-nDNA for up to 28 days (A), from 29 up to 56 days (B) and up to 56 days (C), (D) the diagram illustrating the timeline of blood sample collection.. Each line represents the values for an individual patient, with red indicating an increase over time and blue indicating a decrease over time. Cir-nDNA: circulating nuclear DNA.

### Myeloperoxidase and Neutrophil Elastase assay

Myeloperoxidase (MPO) and Neutrophil Elastase (NE) concentrations were measured using an enzyme-linked immunosorbent assay (ELISA) technique with a Duoset kit from R&D systems. This was used according to the manufacturer’s standard protocol (Duoset R&D Systems, DY008, DY3174, and DY9167-05). Both compounds synergistically contribute to Netosis and especially to DNA decondensation following neutrophil activation, and are specifically anchored to NETs filaments following NETs extracellular release. The healthy cir-nDNA concentration median value for MPO and NE markers (12.8 ng/mL and 7.6 ng/mL, respectively) was determined using a control cohort of 22 healthy individuals (Suppl. Fig. 1). Similar to our definition of a positivity threshold for cir-nDNA, we arbitrarily defined a positivity threshold for MPO and NE concentration (17.0 ng/mL and 10.5 ng/mL, respectively), by adding the standard deviation to the median value of healthy individuals (Suppl. Fig. 1), to evaluate more stringently any discrepancies between abnormal/pathological values and healthy control values. A reproducibility test revealed a coefficient of variation of 3.14% and 5.82% for the quantification of MPO and NE carried out in the same respective plate. A reference sample was added in triplicate in each plate to normalize the value obtained, to address potential variations deriving from manipulator or plate variations. All MPO and NE measurements were carried out using the same batch of plate.

### Statistical analysis

To investigate the associations of patients’ cir-nDNA concentrations at different time points, we utilized the Mann-Whitney test. This was done subsequent to conducting the Shapiro-Wilk test, which confirmed that the sample groups did not adhere to a Gaussian distribution. Spearman correlation evaluates the strength and direction of monotonic associations between two variables. It provides a correlation coefficient to explore the relation between these continuous variables: cir-nDNA, MPO and NE. Our guide for the interpretation of the correlation coefficient is: 0.19, no or negligible relationship; 0.20-0.29, weak but significant relationship if there is a P value <0.05; 0.30 −0.39; moderate but significant relationship if there is a P value <0.05; 0.40-0.69; strong and significant relationship if there is a P value <0.05; and, > 0.70, very strong and significant relationship if there is a P value <0.05. All P values reported are two sided. The significance level was set at 5% (p<0.05); *p < 0.05; **p < 0.01; ***p < 0.001; ****p < 0.0001. Statistical analysis was performed using the Graph Pad Prism 10.0.1 software.

## Results

For this study, we exploited a prospective, multicenter, blinded clinical study (THRuST) which followed up stage III colon cancer patients using an individualized molecular tag selected from a tumor sequencing mutation profile. Plasma samples from 74 patients were included in the study (Suppl.Table 1). In the cohort, 7 patients lacked post-surgery samples, 47 had a single post-surgery sample, and 20 had two post-surgery samples (Fig. 1D). For subsequent analyses comparing pre- and post-surgery samples, paired values from 67 patients were utilized. In cases where two post-surgery samples from a patient fell within the defined timeframe, the most recent sample was prioritized for analysis. Overall, among all blood draws from this cancer cohort, there were 50 (67.6%) and 15 (20.3%) samples which showed higher and twice higher cir-nDNA concentrations than the median for healthy individuals (12.6 +/- 7.2, SD) (Suppl. Table 2 and 3).

Considering all the available 87 post-surgery samples, 77 (88.5%) and 49 (56.3%) plasma samples showed higher and twice higher cir-nDNA concentration than the median for healthy individuals, respectively (Suppl. Table 3-5). Unexpectedly, we observed that 39 out of 67 (58.1%) patients showed two-month post-surgery cir-nDNA amounts higher than their corresponding pre-surgery value. Notably, plasma samples from n 20 (29.9%) and 14 (20.9%) patients out of 67 showed a two-fold and three-fold increase as compared to their pre-surgery values, respectively (Suppl. Table 3A).

Considering that the time to recover from surgery might impact the levels of cir-nDNA, we performed a subgroup analysis of samples by examining post-surgical plasma collected during the first four weeks (days 0-28) separately from those drawn at the later timepoints (days 29-56 since surgery) (Fig.1A-B).

During the 0-28 day post-surgery period, 33 out of 46 (71.7%) patients had a cir-nDNA amount that was higher than their pre-surgery level (Fig. 1A), with 23 out of 46 (50.0%) and 13 out of 46 (28.3%) being 2-fold and 3-fold higher, respectively (Suppl.Fig. 1A, Suppl. Table 3A). Only 8 out of 46 (28.3 %) cases showed post-surgery cir-nDNA levels comparable with those pre-surgery (CV=24%).

During the 28-56 day post-surgery period, 19 out of 37 (51.4%) patients had cir-nDNA levels higher than their pre-surgery levels (Fig.1B), with 6 out of those 37 (28.7%) being 2-fold higher and 3 of 37 (8,11%) being 3-fold higher, respectively (Suppl. Fig. 1). Only 11 out of 37 (29.7%) showed comparable level. The vast majority of cir-nDNA concentrations increased post-surgery, as compared to pre-surgery values. In addition, data showed a trend towards higher elevations in the first four weeks post-surgery (Fig. 1 and Suppl. Fig. 1).

In order to gain further insights into the fluctuations of cir-nDNA levels, we considered the cir-nDNA post-operative values of plasma samples obtained within weekly periods up to 8-weeks post-surgery. Whereas we observed a weak statistical difference between pre-surgery and median cir-nDNA levels in each of the eight weeks subsequent to surgery, as compared to their matched pre-operative values (Fig.2 and detailed in Suppl. Table 2-5), stronger statistical differences were observed between control and post-surgery samples during each week of the eight week post-surgery period we studied (P<0.05 to P<0.0001, Suppl. Table 5). Detailed analysis of data obtained during each of the eight post-surgery weeks is given in Suppl. Information. Generally, the samples analyzed during the first week post-surgery showed the strongest increase in cir-nDNA levels, as compared to subsequent weeks. Those levels showed a tendency to gradually decrease week on week up to the fourth week post-surgery, while remaining elevated in the subsequent weeks up to the eighth post-surgery week (100%, 90.9%, 81.8%, 73.3%, 58.8%, 72.7% and 62.5% in the 1-, 2-, 3-, 4-, 5-, 6-, 7-days post-surgery week, respectively) (Suppl. Fig.1A and Suppl. Table 2). Post-surgery cir-nDNA concentration increase as compared to pre-surgery appears to stabilize from the fifth to the eighth week post-surgery (58.8% to 72.7%). In addition, the median values obtained pre-surgery or during the first seven weeks post-surgery are all statistically higher than the median of healthy individuals (P<0.05 to P<0.0001, Suppl. Table 6). It is also significant that all median values obtained post-surgery for each week are greater than the medians obtained pre-surgery (fig. 3A and Suppl. Table 6). It should be noted that post-surgery cir-nDNA levels are statistically higher than pre-surgery levels in only the first and second week post-surgery (P<0.0001, Suppl. Table 6).

**Figure 2:**
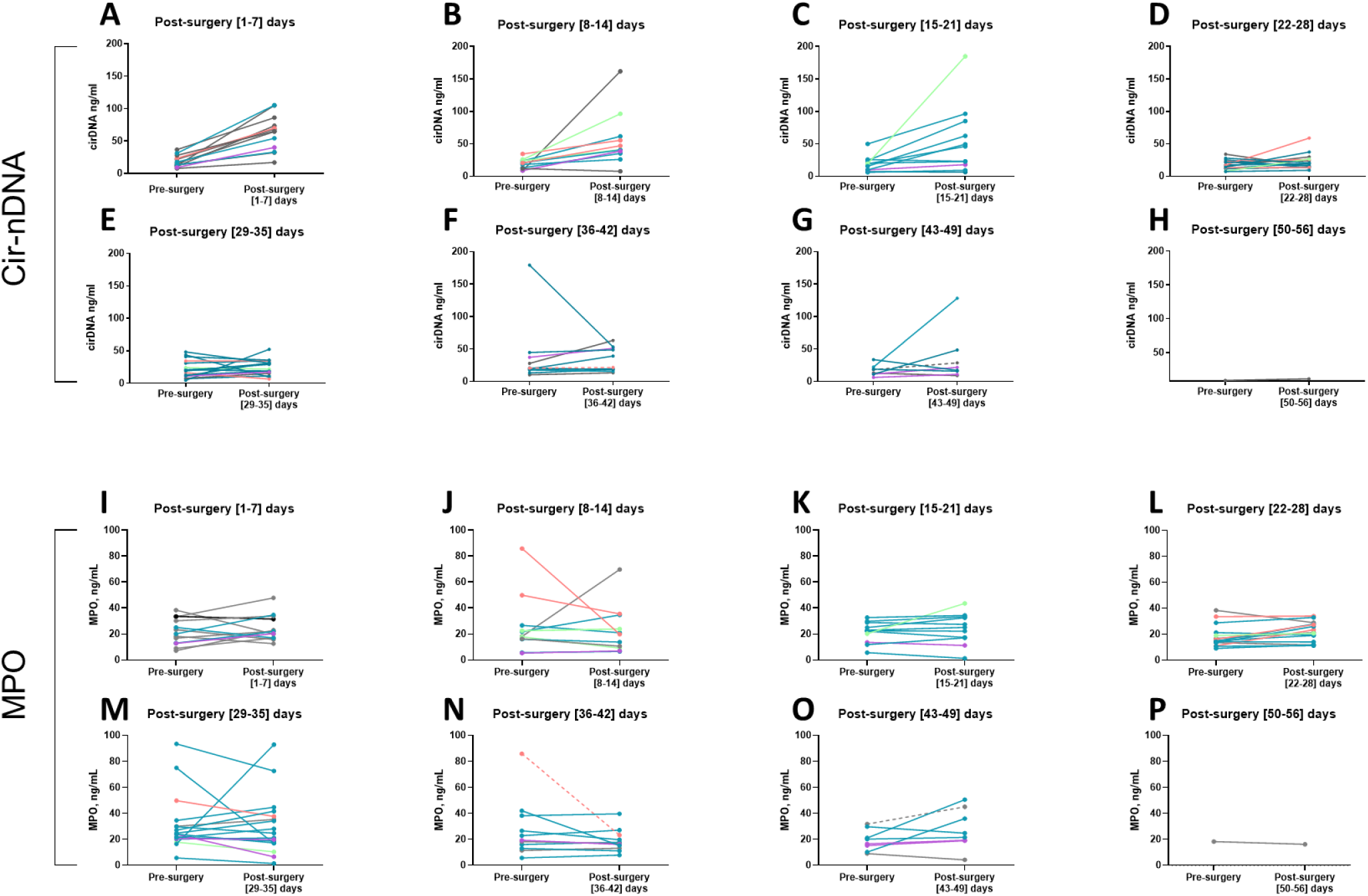
Comparative analysis of cir-nDNA and MPO concentrations in pre-surgery and every week in post-surgery. The graphs representing per week the comparative analysis post-surgery versus the pre-surgery values. The analysis of Cir-nDNA is showed for the first (A), second (B), third (C), fourth (D), fifth (E), sixth (F), seventh(G), eighth (H) week after surgery. The analysis of MPO showed for the first (I), second (J), third (K), fourth (L), fifth (M), sixth (N), seventh(O), eighth (P) week after surgery. Violet (category 1) - patients with no relapse or adverse event whose marker values return to healthy levels after 2 months; bleu (category 2) - patients with no relapse or adverse event whose marker values do not return to healthy levels after 2 months; green (category 3) - patients with relapse; rose (category 4) - patients with adverse events; gray (category 5) - patients without relapse or adverse events with less than 2 months follow up; gray dashed line - patients who got chemotherapy before sampling. Cir-nDNA: circulating nuclear DNA; MPO – myeloperoxidase.

**Figure 3:**
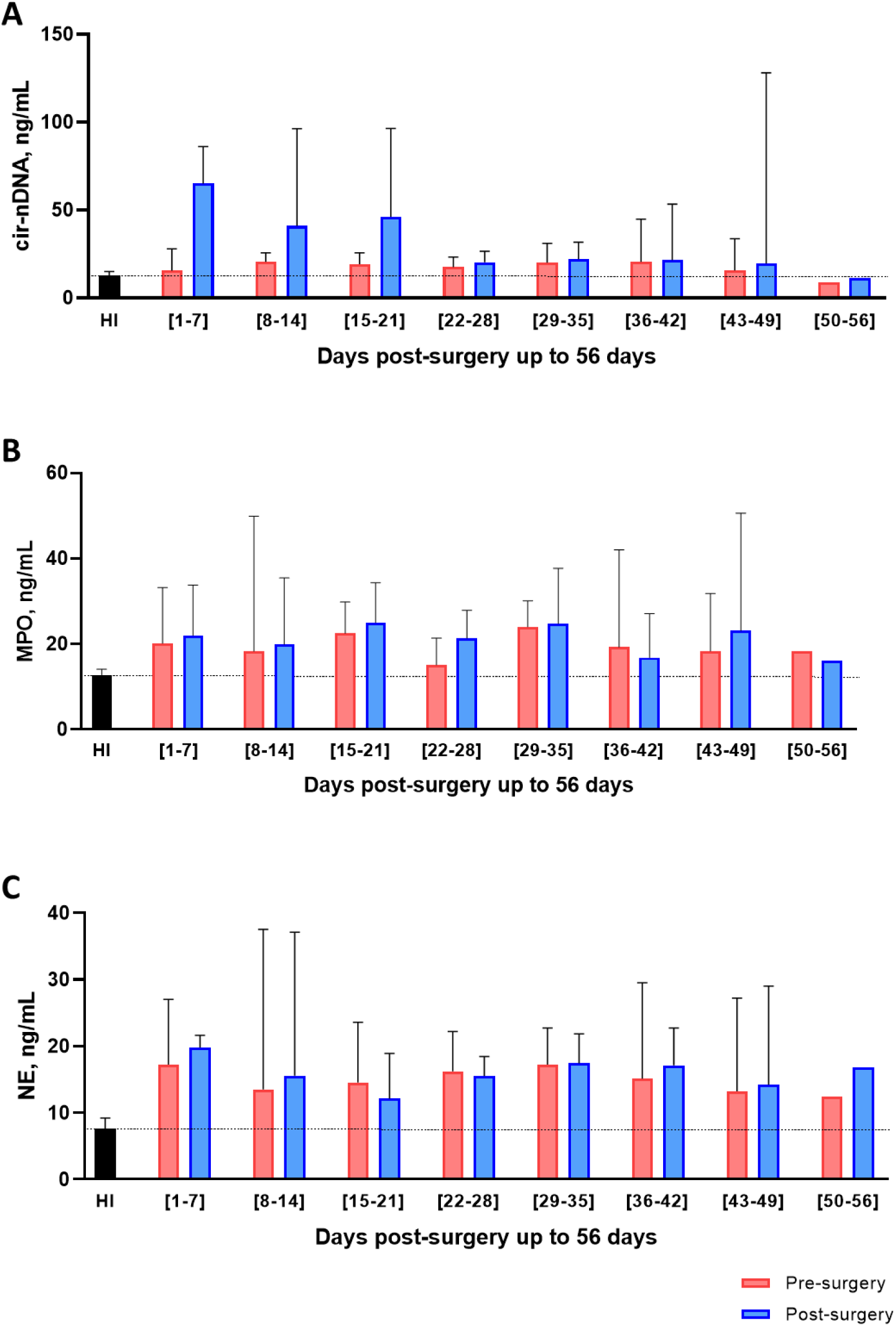
Comparative analysis of cir-nDNA and MPO concentrations in pre-surgery and every week in post-surgery versus healthy individuals. The graphs representing the comparative analysis circulating DNA (A), MPO (B) and NE (C) concentrations obtained post-surgery are compared with the values obtained pre-surgery and with the median obtained in the cohort of healthy individuals (HI). The bar plots indicated in median with 95% CI. Cir-nDNA: circulating nuclear DNA; MPO – myeloperoxidase; NE - neutrophil elastase.

No strong trend emerges from the results when the plasmas originated from: (1), patients without relapse or adverse event whose marker values return at two months post-surgery to levels equivalent to those of healthy individuals; (2), patients without relapse or adverse event whose marker values at two months post-surgery do not return to the levels of healthy controls; (3), patients experiencing a relapse or adverse event; and (4), patients without relapse or adverse event with less than two months of post-surgery follow-up (Fig.2A-H). Note, the plasmas of the two patients who received chemotherapy before their post-surgery blood sample was obtained showed a relative stability compared to their pre-surgery values.

### Follow-up of the post-surgery values of the NETs markers

The changes in MPO plasma concentrations as assessed in weekly intervals up to 8 weeks post-surgery as a function of the pre-surgery values is presented in Fig. 2. MPO concentration values in plasma samples taken during the first week post-surgery increased in comparison to the baseline, with only two samples showing values lower than pre-surgery. The MPO concentrations in samples drawn during the second week showed distinctly varied trends, with both strong increases and decreases in value, as compared to pre-surgery values. The MPO concentration values obtained during the third and fourth week (15 to 28 days post-surgery) are relatively stable compared to the pre-surgery levels. Post-surgery MPO concentrations appear to be mostly higher than those observed pre-surgery except post-surgery values obtained from the fifth to the eighth weeks that remain relatively stable (Suppl. Table 2 and 5). Five plasmas showed very strong variations, both positive and negative (Fig. 3B and 4A, Suppl. Table 4B).

**Figure 4:**
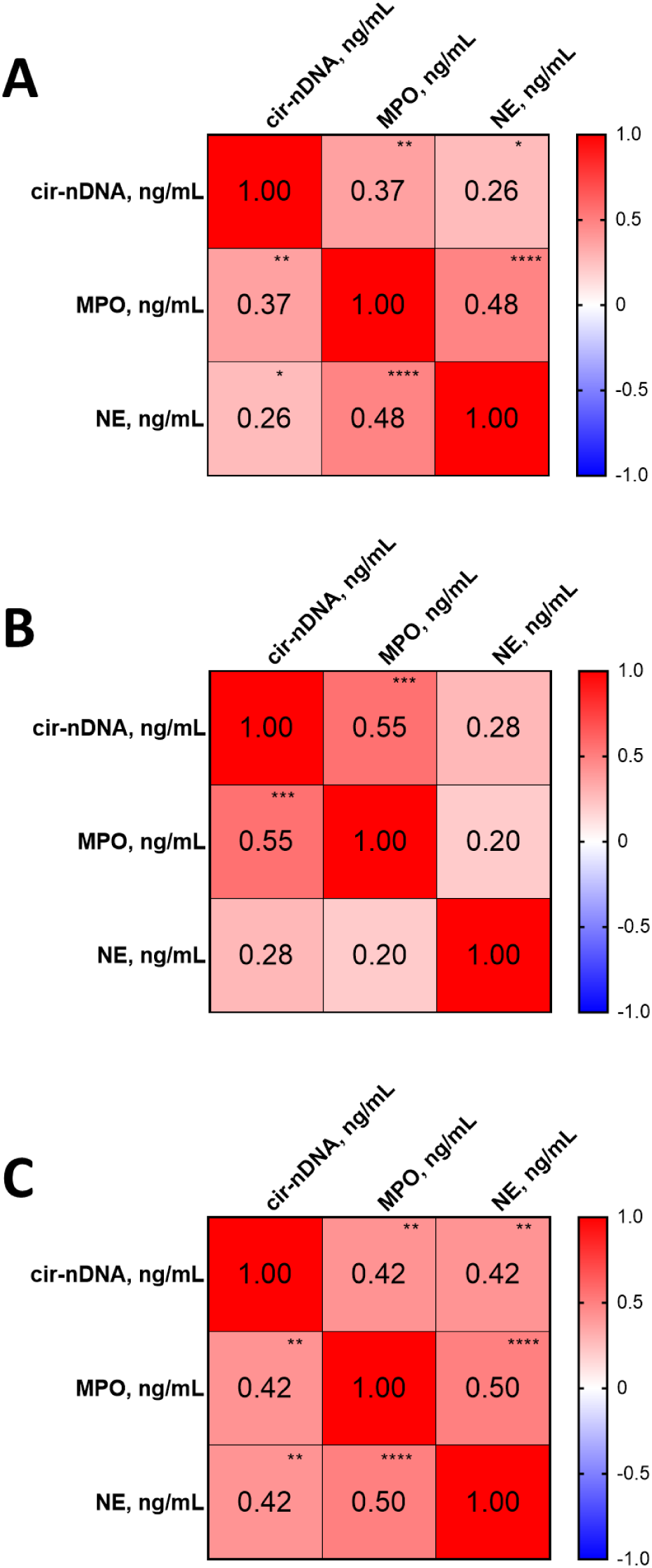
Correlation analysis of cir-nDNA, MPO and NE concentrations in healthy individuals, pre-surgery and post-surgery for patients. Spearman correlation study between all MPO, NE and cir-nDNA values. (A), values of healthy individuals HI (n=22); Spearman P-value associating MPO and NE, MPO and cir-DNA and NE and cir-nDNA are 0.007, 0.118, 0.554; (B), pre-surgery values (n=74). Spearman P-value associating MPO and NE, MPO and cir-DNA and NE and cir-nDNA are 1.60 x 10^-5^, 1.36 x 10^-3^ and 0.022; (C), 1-21 days of follow-up values (n=35). Spearman P-value associating MPO and NE, MPO and cir-DNA and NE and cir-nDNA are 0.246, 5.71 x 10^-4^ and 0.110; (D), 22-56 days of follow-up values (n=52). Spearman P-value associating MPO and NE, MPO and cir-DNA and NE and cir-nDNA are 1.76 x 10^-4^, 1.94 x 10^-3^ and 2.03 x 10^-3^, respectively. Cir-nDNA: circulating nuclear DNA; MPO: myeloperoxidase.

Altogether, all MPO values obtained pre-surgery or during the first eight weeks post-surgery are statistically higher than the median in healthy individuals (Fig.3B). Except during the sixth week post-surgery, all post-surgery values were found to be higher but not statistically different from the values obtained pre-surgery. All NE concentrations determined pre-surgery or during the first eight weeks post-surgery are statistically higher than the median of healthy individuals (Suppl. Table 4C). There is no significant difference between the concentrations determined for each week up to the eighth week post-surgery (Fig.4C, Suppl. Fig.1B and C, and Suppl. Table 4C).

Analysis of the association of the NETs markers (MPO and NE) and total circulating DNA were performed using the Spearman correlation matrix (Fig. 4). As previously observed, no association between cir-nDNA and NETs markers was found in plasma from healthy individuals, whereas a significant association exists between NE and MPO, which gives rise to their constitutive expression, albeit low level of plasma amount present in healthy individuals (Fig. 4A). We observed a very strong association of the three markers (cir-nDNA, MPO and NE) during the first three weeks post-surgery, and a slightly weaker association during the following five weeks (Fig. 4B and C).

## Discussion

This is an ancillary study of the THRuST clinical study on stage III colon cancer. It focuses on the variation of cir-nDNA levels within the first eight weeks, in order to align with the timeframe within which the clinician must decide the type and the length of adjuvant therapy, guided by the detection of the MRD as determined by cir-nDNA analysis.

As expected, pre-surgery cir-nDNA concentrations are elevated by comparison to the HI median (12), with high levels being observed up to two weeks post-surgery (13). Unexpectedly, we observed that generally post-surgery cir-nDNA concentrations do not decrease up to eight weeks post-surgery, as compared to the median of healthy individuals and to the matched pre-surgery levels. The differences between pre- and post-surgery levels are greater in the first two weeks post-surgery, and particularly so within the first week. In addition, cir-nDNA levels may greatly increase in some patients, up to 18-fold (Suppl. Table 5). Only 18.9% of patient samples showed a significant decrease in cir-nDNA levels in the eight weeks post-surgery, whereas it might have been assumed that the trauma consequences of surgery (inflammation, cell death, et-cetera) would not impact cir-nDNA concentration beyond the first month post-surgery. Almost all cir-nDNA concentrations as determined in the 8 weeks post-surgery were higher than pre-surgery levels, and did not decrease following surgery in cases where no relapse occurred. This finding challenges previous paradigms (14): While the vast majority of the authors in this field have speculated that cir-mutDNA as well as cir-nDNA concentrations would decline in the case of curative treatment, it should be noted that a small number of previous reports have shown data consistent with our observations (15). We and others (8)(16) have previously inferred that the surgical procedure would enhance the release of cir-nDNA and consequently arbitrarily did not collect blood before two weeks surgery (8), (17).

Two potential physiological effects linked to surgery may explain our observations: (i), “Surgical trauma” denotes the physical damage inflicted on tissues during surgery, encompassing incisions, tissue manipulation, and organ removal, leading to local inflammation and tissue healing; and (ii), “Surgical stress” refers to the physiological and psychological response engendered by the stressors associated with surgery, including the release of stress hormones and systemic changes in heart rate and immune function (18). While these two phenomena are related, they represent different aspects of the body’s reaction to surgical procedures. Given that cir-nDNA levels can remain elevated for up to two months post-surgery, we assume that both phenomena might persist during the two months post-operative period for stage III colon cancer patients.

Given that previous literature has linked an innate immune response involving an inflammatory process, namely the formation of NETs (Netosis), with cancer progression (19) (20)(21) as well as with cir-nDNA release(10), we analysed the presence of NETs markers (MPO and NE) within the same plasma samples in which cir-nDNA content was assessed in this study. NETs are scaffolds of DNA associated with various molecules, such as cytotoxic enzymes (MPO) or proteases (NE), which are released into the extracellular milieu (22). Their primary role is to control the spread of microbes during the first hours of infection. However, dysregulated NETs production may lead to deleterious pathophysiological effects (22). Our data clearly demonstrates that cir-nDNA levels are strongly associated with NETs markers from the first to the eight-week post-surgery. Neither of these observations has been reported previously, and as such they provide a new paradigm of both cir-nDNA origin and the post-surgery follow-up of stage III colon cancer patients. This observation relies on our recent observations of stage II-III CRC and prostate cancer patients in whom this association was revealed in both the immediate peri-surgery period (1-72 hours post-surgery), and in follow-up up to two years post-surgery, for the same patients as those considered in this study (23).

Historically, investigations regarding the potential of cir-nDNA concentration as a single biomarker in oncology have tended to take place in the context of evaluations of treatment response in metastatic disease or of the presence of recurrent cancer post-surgery, or of the association of post-surgery recurrence with overall survival (24). Nevertheless, in patients with cancer, it may be assumed that cir-nDNA plasma concentration measured in ng/mL will range from just a few to several thousand, which overlaps with the 1-20 ng/mL concentration range for healthy individuals (12)(25). Applying standardization and guidelines, Meddeb et al (12) compared the cir-nDNA concentration of healthy individuals (4 ng/mL, median; 0.4-22 ng/mL, range) and mCRC patients at diagnosis (13 ng/mL, median; 1-350 ng/mL, range). Because of its relatively low performance, total cir-nDNA analysis, was not at that point considered as a single biomarker. Indeed, numerous investigators have observed the progressive decrease in levels of circulating mutant DNA (cir-mutDNA) during the follow-up period of tumour-free patients(4).

The significance of NETs as a principal source of cir-nDNA in healthy and cancer individuals has recently been directly proven (11) and is indirectly supported by cir-nDNA fragmentome and methylome studies. In a milestone report(26), Shendure’s team investigated the nucleosome occupancy as determined by fragmentomics, and revealed that lymphoid or myeloid origins have the largest proportions consistent with hematopoietic cells as the dominant source of cir-nDNA in healthy individuals, in contrast to patients with cancer. The work by Dor’s team (27) on cir-nDNA methylome from HI also showed that white blood cells are among the main cir-nDNA cells-of-origin (∼85%), with granulocytes and erythrocyte progenitors being prominent amongst these. Recent reports have used methylome deconvolution to further characterize the neutrophil origin as one of the most important among a great variety of cells-of-origin (27), thus opening up a wide range of opportunities for diagnosis and for the interrogation of the process of tumour progression. While lower proportions of cir-nDNA of neutrophil origin have been detected in patients with cancer than in healthy individuals in normal conditions (27), of all the different sources of cir-nDNA in patients with cancer, the neutrophil origin appears to be amongst the most important. A link has been made between inflammation, NETs and cir-nDNA release in infectious diseases and sterile inflammatory diseases (22), such as in cancer (4)(10). One possible explanation of this could be that NETs are produced from circulating neutrophils as well as from neutrophils on within the tumor-environment mass.

In addition, the formation of NETs for the post-surgery determination of cir-nDNA concentration seems to lead to high inter-individual variation as observed here among stage III colon cancer patients, which ranges up to 11.3 and 18-fold during the first and the second month, respectively.

The patient number of the patient cohort (N=74) does not allow to statistically evaluate whether cir-nDNA or NETs markers are of value in regards to the prognosis of the recurrence or adverse events given the low proportion of relapse (Fig. 2 and Suppl. Table 7) and adverse events (Fig. 2 and Suppl. Table 8, 14.9% and 6.8%, respectively). Another limitation of the study is the lack of the possibility, so far, to explain the inter-individual high variability of the NETs formation. Thus, use of a much larger cohort and detailed analysis of the individual physiological and genetic/epigenetic factors are necessary.

We believe it would be misleading to define an optimal post-surgery blood collection time for MRD detection. At best, we estimate that the time range within which the cir-nDNA content of collected blood is likely to be the lowest, and in which there will probably by the least inter-individual variability, is between the fourth and the sixth week post-surgery (Suppl. Fig. 1A).

Our observations appear to cast considerable doubt on the potential of post-surgery detection of MRD guided by cir-mutDNA analysis, in particular in colorectal cancer (CRC) patient management care. Thus, Tie at al’s studies (9), for instance, proved that MRD detection through the use of cir-nDNA analysis would greatly help the stratification of patients with regard to adjuvant therapy intensity. Given that these studies are based on the detection of cir-nDNA bearing a mutation which shows the highest mutation allele frequency (MAF, proportion of mutant DNA among total DNA) as determined from blood samples by NGS, selection of the analysis of cir-mutDNA bearing such a mutation may not always be reliable with respect to the level of WT cir-nDNA, which may vary according to the patient. Given the low percentage of mutant cell clone within the tumor which can sometimes occur, the sensitivity of the detection is a critical issue with respect to MAF determination, due to the method’s limit of detection (LOD) (from 0.003% to 0.1%, using IntPlex qPCR and conventional NGS) (15). Consequently, the selection of the mutation to be tracked post-surgery, along with its monitoring from cir-mutDNA based on MAF, are sub-optimal. Furthermore, the doubtful reliability of using MAF from plasma is a factor to be considered, for instance in the use of a MAF threshold for selecting patients for targeted therapy according to tumor mutation status (16), (28).

Our observations point to a need to reassess the criteria for MRD blood collection time and for the use of MAF/VAF. Indeed, cir-nDNA MAF should be considered a “false friend” marker when assessing the proportion of malignant cells within a tumor. To these concerns, our data support the following recommendations: (i), for MRD detection, precise measurement of cir-nDNA concentration must be carried out with appropriate methods; (ii), in the case of high cir-nDNA concentrations (over a threshold, i.e. ∼2-fold control HI concentration), on one hand MPO and/or NE plasma concentration must be evaluated, and on the another hand MPO and/or NE plasma concentration must be evaluated, and in another hand the most sensitive method available must be used to assess MAF; (iii), the absolute mutation quantification (concentration of cir-mutDNA) must be used, rather than the relative MAF; and (iv), in regards to the selection of patients towards targeted therapy the use of a MAF-based threshold must not be used when determining tumour mutation status.

## Ethical approval statement

Informed consent was obtained from all individuals and/or caregivers, and all clinical procedures and genetic testing, including data collection and report, were in accordance with the declaration of Helsinki and approved by the local ethical committees or followed other local guidelines: protocol 015-FP018 by the Ethics Committee at the Hospital Universitari Bellvitge, protocol ID CE IRCCS n.233/2018 approved by the Ethical Committee of the Candiolo Cancer Institute FPO IRCCS, protocol 2019/34 approved by CPP Ouest II; protocol PR(AG)235/2018 approved by Vall d’Hebron Ethical Committee.

## DATA Sharing Statement

The data that support these findings of the study are available upon request from the corresponding authors.

## Data Availability

All data produced in the present study are available upon reasonable request to the authors

## Acknowledgments

The authors thank the excellent technical assistance of F. Frayssinoux (IRCM, Institut de Recherche en Cancérologie de Montpellier, INSERM U1194, Université de Montpellier, Institut régional du Cancer de Montpellier, Montpellier, F-34134298, France) and Cormac Mc Carthy (Mc Carthy Consultant, Montpellier) for English editing (financial compensation). The authors thank K. Bibova from the TRANSCAN and the Dpt of International coopfolderation, Slovak Academy of Sciences. VHIO would like to acknowledge the Cellex Foundation for providing research facilities and equipment, and the FERO Foundation for their funding support. Authors acknowledge financial support by the Instituto de Salud Carlos III-Investigación en Salud (AC17/00119), the Fundación AECC (CLSEN19001ELEZ), and Generalitat de Catalunya (Agència de Gestió d’Ajuts Universitaris i de Recerca, AGAUR 2021SGR01567 We thank the healthy donors who participated in this study. We thank all patients and their families participated in this study for their trust.

## Funding

This work was supported by the European ERA-NET grant on Translational Cancer Research (TRANSCAN-2) “Minimally and non-invasive methods for early detection and/or progression of cancer”. This work was partially supported by SIRIC Montpellier Cancer Grant INCa_Inserm_DGOS_12553,by MSD AVENIR [MSD-THRuST grant and by the Société Française des Acides Nucléiques Circulants (SFAC). This work was supported in Spain by Fundación Científica Asociación Española Contra el Cancer (FC-AECC) / Instituto de Salud Carlos III (ISCIII). This work was also supported in Italy by the Italian Ministry of Health.

---

**Supplementary figure 1.**
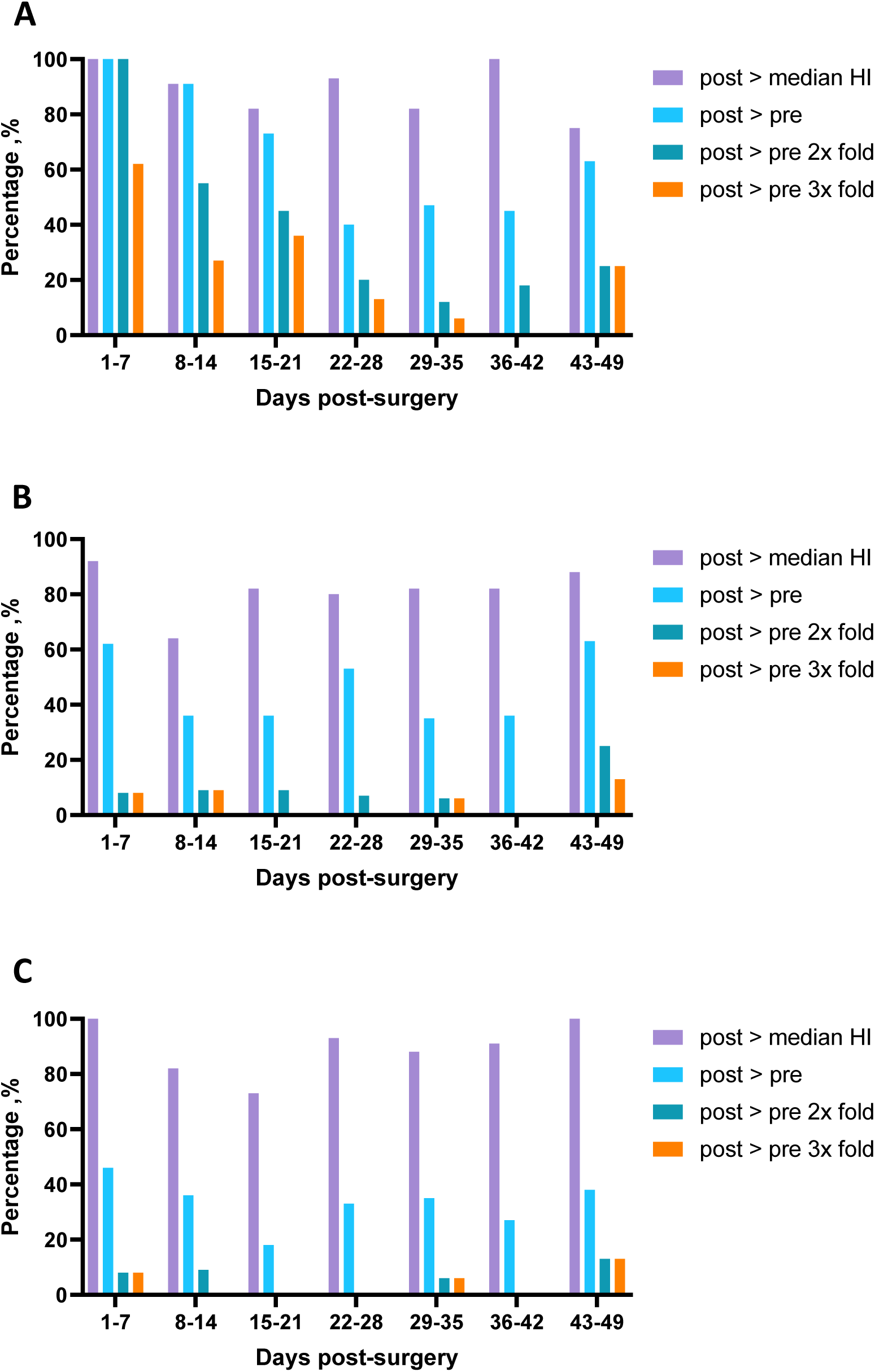
Proportion of patients showing greater post-surgery values as compared to healthy individuals (HI) median and pre-surgery values in respect to the concentration of circulating nuclear DNA (A), myeloperoxidase (B) and neutrophils elastase (C).

## Notes

### Competing Interest Statement

The authors have declared no competing interest.

### Funding Statement

This study was funded by the European ERA-NET grant on Translational Cancer Research (TRANSCAN-2). This study was funded by SIRIC Montpellier Cancer Grant INCa_Inserm_DGOS_12553,by MSD AVENIR [MSD-THRuST grant and by the Societe Francaise des Acides Nucleiques Circulants (SFAC). This study was funded by Spain by Fundacion Cientifica Asociacion Espanola Contra el Cancer (FC -AECC) / Instituto de Salud Carlos III (ISCIII). This study was funded by the Italian Ministry of Health.

### Author Declarations

Ethics Committee of Candiolo Cancer Institute FPO-IRCCS gave ethical approval for this work. Ethics Committee at the Hospital Universitari Bellvitge gave ethical approval for this work. Ethics Committee CPP Ouest II of ICM gave ethical approval for this work.Ethics Committee Vall Hebron Ethical Committee gave ethical approval for this work

## References

1. André T, Meyerhardt J, Iveson T, Sobrero A, Yoshino T, Souglakos I, et al. Effect of duration of adjuvant chemotherapy for patients with stage III colon cancer (IDEA collaboration): final results from a prospective, pooled analysis of six randomised, phase 3 trials. The Lancet Oncology. 2020;21:1620–9.

2. Taieb J, Basile D, Seligmann J, Argiles G, André T, Gallois C, et al. Standardizing data collection in adjuvant colon cancer trials: A consensus project from the IDEA and ACCENT international consortia and national experts. Eur J Cancer. 2024;206:114118.

3. Dong Q, Chen C, Hu Y, Zhang W, Yang X, Qi Y, et al. Clinical application of molecular residual disease detection by circulation tumor DNA in solid cancers and a comparison of technologies: review article. Cancer Biology & Therapy. 2023;24:2274123.

4. Conca V, Ciracì P, Boccaccio C, Minelli A, Antoniotti C, Cremolini C. Waiting for the “liquid revolution” in the adjuvant treatment of colon cancer patients: a review of ongoing trials. Cancer Treatment Reviews. 2024;126:102735.

5. Bent A, Kopetz S. Going with the Flow: The Promise of Plasma-Only Circulating Tumor DNA Assays. Clin Cancer Res. 2021;27:5449–51.

6. Guo N, Zhou Q, Chen X, Zeng B, Wu S, Zeng H, et al. Circulating tumor DNA as prognostic markers of relapsed breast cancer: a systematic review and meta-analysis. Journal of the National Cancer Center. 2024;4:63–73.

7. Bronkhorst AJ, Ungerer V, Diehl F, Anker P, Dor Y, Fleischhacker M, et al. Towards systematic nomenclature for cell-free DNA. Hum Genet [Internet]. 2020 [cited 2021 Feb 15]; Available from: http://link.springer.com/10.1007/s00439-020-02227-2

8. Thierry AR, Mouliere F, El Messaoudi S, Mollevi C, Lopez-Crapez E, Rolet F, et al. Clinical validation of the detection of KRAS and BRAF mutations from circulating tumor DNA. Nat Med. 2014;20:430–5.

9. Tie J, Kinde I, Wang Y, Wong HL, Roebert J, Christie M, et al. Circulating tumor DNA as an early marker of therapeutic response in patients with metastatic colorectal cancer. Ann Oncol. 2015;26:1715–22.

10. Pastor B, Abraham J-D, Pisareva E, Sanchez C, Kudriavstev A, Tanos R, et al. Association of neutrophil extracellular traps with the production of circulating DNA in patients with colorectal cancer. iScience. 2022;25:103826.

11. Pisareva E, Mihalovičová L, Pastor B, Kudriavstev A, Mirandola A, Mazard T, et al. Neutrophil extracellular traps have auto-catabolic activity and produce mononucleosome-associated circulating DNA [Internet]. Molecular Biology; 2022 Sep. Available from: http://biorxiv.org/lookup/doi/10.1101/2022.09.01.506266

12. Meddeb R, Dache ZAA, Thezenas S, Otandault A, Tanos R, Pastor B, et al. Quantifying circulating cell-free DNA in humans. Sci Rep. 2019;9:5220.

13. Diehl F, Li M, Dressman D, He Y, Shen D, Szabo S, et al. Detection and quantification of mutations in the plasma of patients with colorectal tumors. Proc Natl Acad Sci U S A. 2005;102:16368–73.

14. Thierry AR, Pisareva E. A New Paradigm of the Origins of Circulating DNA in Patients with Cancer. Cancer Discovery. 2023;13:2122–4.

15. Bos MK, Nasserinejad K, Jansen MPHM, Angus L, Atmodimedjo PN, de Jonge E, et al. Comparison of variant allele frequency and number of mutant molecules as units of measurement for circulating tumor DNA. Mol Oncol. 2021;15:57–66.

16. Henriksen TV, Reinert T, Christensen E, Sethi H, Birkenkamp-Demtröder K, Gögenur M, et al. The effect of surgical trauma on circulating free DNA levels in cancer patients— implications for studies of circulating tumor DNA. Molecular Oncology. 2020;14:1670–9.

17. Cohen SA, Kasi PM, Aushev VN, Hanna DL, Botta GP, Sharif S, et al. Kinetics of postoperative circulating cell-free DNA and impact on minimal residual disease detection rates in patients with resected stage I-III colorectal cancer. JCO. 2023;41:5–5.

18. Rosen AW, Gögenur M, Paulsen IW, Olsen J, Eiholm S, Kirkeby LT, et al. Perioperative changes in cell-free DNA for patients undergoing surgery for colon cancer. BMC Gastroenterol. 2022;22:168.

19. Park J, Wysocki RW, Amoozgar Z, Maiorino L, Fein MR, Jorns J, et al. Cancer cells induce metastasis-supporting neutrophil extracellular DNA traps. Sci Transl Med. 2016;8:361ra138.

20. Mousset A, Lecorgne E, Bourget I, Lopez P, Jenovai K, Cherfils-Vicini J, et al. Neutrophil extracellular traps formed during chemotherapy confer treatment resistance via TGF-β activation. Cancer Cell. 2023;41:757–775.e10.

21. Bisanzi S, Puliti D, Picozzi G, Romei C, Pistelli F, Deliperi A, et al. Baseline Cell-Free DNA Can Predict Malignancy of Nodules Observed in the ITALUNG Screening Trial. Cancers (Basel). 2024;16:2276.

22. Papayannopoulos V. Neutrophil extracellular traps in immunity and disease. Nat Rev Immunol. 2018;18:134–47.

23. Kudriavtsev A, Pastor B, Mirandola A, Pisareva E, Gricourt Y, Capdevila X, et al. Association of the immediate perioperative dynamics of circulating DNA levels and neutrophil extracellular traps formation in cancer patients. Precision Clinical Medicine. 2024;7:pbae008.

24. Mehra N, Dolling D, Sumanasuriya S, Christova R, Pope L, Carreira S, et al. Plasma Cell-free DNA Concentration and Outcomes from Taxane Therapy in Metastatic Castration-resistant Prostate Cancer from Two Phase III Trials (FIRSTANA and PROSELICA). European Urology. 2018;74:283–91.

25. Phallen J, Sausen M, Adleff V, Leal A, Hruban C, White J, et al. Direct detection of early-stage cancers using circulating tumor DNA. Sci Transl Med. 2017;9:eaan2415.

26. Snyder MW, Kircher M, Hill AJ, Daza RM, Shendure J. Cell-free DNA Comprises an In Vivo Nucleosome Footprint that Informs Its Tissues-Of-Origin. Cell. 2016;164:57–68.

27. Moss J, Magenheim J, Neiman D, Zemmour H, Loyfer N, Korach A, et al. Comprehensive human cell-type methylation atlas reveals origins of circulating cell-free DNA in health and disease. Nat Commun. 2018;9:5068.

28. Urbini M, Marisi G, Azzali I, Bartolini G, Chiadini E, Capelli L, et al. Dynamic Monitoring of Circulating Tumor DNA in Patients With Metastatic Colorectal Cancer. JCO Precision Oncology. 2023;e2200694.

