## Supplementary table for "Post-surgery level of circulating DNA in stage III colon cancer patients: impact on the reliability of minimal residual disease detection"

Supplementary Table 1

| **ID patient** | **Blood collection time (Day)** | **cir-nDNA, ng/mL** | **MPO, ng/mL** | **NE, ng/mL** |
| --- | --- | --- | --- | --- |
| 01-002 | 0 | 15.50 | 16.9 | 16.5 |
|  | 4 | 64.60 | 22.3 | 19.8 |
|  | 27 | 29.90 | 21.4 | 15.1 |
| 01-003 | 0 | 7.28 | 28.77 | 12.95 |
|  | 27 | 9.20 | 32.84 | 13.53 |
| 01-006 | 0 | 9.90 | 32.65 | 27.35 |
|  | 21 | 49.34 | 34.32 | 20.28 |
| 01-011 | 0 | 7.80 | 30.1 | 12.2 |
|  | 4 | 16.90 | 33.8 | 8.7 |
|  | 34 | 15.20 | 35.4 | 17.4 |
| 01-021 | 0 | 10.90 | 33.2 | 23.2 |
|  | 4 | 73.60 | 47.8 | 26.7 |
| 01-023 | 0 | 21.70 | 38.4 | 22.2 |
|  | 4 | 68.30 | 20.0 | 20.2 |
|  | 24 | 23.90 | 28.9 | 17.4 |
| 01-029 | 0 | 23.20 | 33.55 | 16.23 |
|  | 4 | 69.90 | 31.43 | 18.26 |
|  | 28 | 25.80 | 33.94 | 18.45 |
| 01-033 | -3 | 18.24 | 31.82 | 24.43 |
|  | 45 | 28.83 | 45.12 | 29.03 |
| 01-035 | 0 | 48.43 | 27.10 | 13.97 |
|  | 35 | 31.04 | 41.68 | 13.67 |
| 01-036 | 0 | 13.20 | 25.04 | 35.78 |
|  | 4 | 32.88 | 17.05 | 31.98 |
| 01-048 | 0 | 19.13 | 21.35 | 12.30 |
|  | 31 | 30.70 | 28.13 | 16.43 |
| 01-050 | -1 | 9.30 | 12.9 | 5.7 |
|  | 3 | 40.00 | 20.4 | 18.1 |
| 01-053 | 0 | 179.27 | 42.00 | 15.10 |
|  | 42 | 53.31 | 15.29 | 7.47 |
| 01-054 | 0 | 18.93 | 20.172 | 18.68 |
|  | 45 | 15.94 | 21.505 | 16.85 |
| 01-070 | 0 | 6.33 | 16.567 | 12.74 |
|  | 45 | 10.89 | 19.310 | 8.82 |
| 01-077 | 0 | 34.00 | 13.64 | 19.87 |
|  | 28 | 15.34 | 11.68 | 3.84 |
| 01-081 | 0 | 22.22 | 10.17 | 11.03 |
|  | 49 | 128.02 | 36.06 | 15.95 |
| 01-087 | 0 | 11.42 | 15.28 | 13.71 |
|  | 49 | 21.74 | 18.92 | 11.62 |
| 01-092 | 0 | 20.90 | 26.6 | 21.7 |
|  | 9 | 40.50 | 21.0 | 20.0 |
|  | 37 | 18.10 | 19.7 | 18.8 |
| 01-093 | 0 | 36.80 | 23.4 | 27.0 |
|  | 5 | 86.00 | 15.9 | 11.3 |
| 01-101 | 0 | 11.34 | 14.71 | 13.24 |
|  | 28 | 14.03 | 27.88 | 15.42 |
| 01-105 | 0 | 44.77 | 38.276 | 21.18 |
|  | 38 | 48.77 | 39.687 | 20.53 |
| 01-108 | 0 | 11.86 | 16.41 | 10.44 |
|  | 8 | 7.62 | 10.68 | 4.05 |
| 01-123 | -3 | 10.23 | 11.46 | 8.62 |
|  | 36 | 13.47 | 13.21 | 8.69 |
| 01-124 | 0 | 21.90 | 9.15 | 11.05 |
|  | 23 | 16.07 | 11.96 | 8.48 |
| 01-125 | 0 | 16.33 | 23.94 | 12.40 |
|  | 34 | 6.64 | 6.54 | 2.21 |
| 01-130 | 0 | 31.10 | 20.2 | 17.2 |
|  | 4 | 105.20 | 34.7 | 14.7 |
|  | 30 | 34.80 | 21.1 | 20.0 |
| 01-136 | 0 | 18.30 | 16.0 | 29.5 |
|  | 13 | 26.00 | 13.9 | 38.0 |
|  | 37 | 19.50 | 17.8 | 22.7 |
| 01-139 | 0 | 27.90 | 18.3 | 24.3 |
|  | 4 | 65.00 | 12.6 | 21.1 |
|  | 41 | 63.10 | 16.8 | 25.0 |
| 01-141 | -15 | 25.40 | 21.4 | 30.6 |
|  | 28 | 13.10 | 19.5 | 26.5 |
| 01-144 | 0 | 24.00 | 17.9 | 31.2 |
|  | 8 | 40.90 | 9.2 | 22.5 |
|  | 35 | 22.00 | 10.4 | 26.4 |
| 01-150 | 0 | 23.70 | 21.87 | 11.65 |
|  | 10 | 61.60 | 34.66 | 7.16 |
|  | 30 | 17.40 | 18.73 | 11.52 |
| 01-157 | 0 | 17.60 | 13.0 | 14.5 |
|  | 4 | 54.20 | 22.0 | 21.6 |
|  | 38 | 15.60 | 11.1 | 17.1 |
| 01-168 | 0 | 7.00 | 5.7 | 9.3 |
|  | 19 | 6.40 | 1.3 | 7.1 |
|  | 34 | 11.40 | 1.4 | 11.6 |

Supplementary Table 1 - continued

| **ID patient** | **Blood collection time (Day)** | **cir-nDNA. ng/mL** | **MPO. ng/mL** | **NE. ng/mL** |
| --- | --- | --- | --- | --- |
| 01-169 | 0 | 9.00 | 18.3 | 12.4 |
|  | 12 | 161.90 | 69.7 | 37.1 |
|  | 50 | 11.40 | 16.2 | 16.9 |
| 01-171 | 0 | 11.90 | 7.3 | 13.2 |
|  | 4 | 104.90 | 22.9 | 20.9 |
| 01-172 | 0 | 13.10 | 9.0 | 27.2 |
|  | 3 | 32.20 | 17.0 | 16.4 |
|  | 48 | 9.10 | 4.2 | 12.5 |
| 01-178 | 0 | 13.10 | 5.7 | 11.1 |
|  | 9 | 35.20 | 6.6 | 12.2 |
|  | 37 | 18.60 | 7.9 | 12.4 |
| 01-183 | 0 | 17.60 | 15.1 | 22.4 |
|  | 28 | 18.90 | 19.1 | 15.6 |
| 02-001 | 33 | 35.74 | 72.602 | 24.14 |
|  | -23 | 41.00 | 93.527 | 30.47 |
| 02-002 | -28 | 43.67 | 75.01 | 53.46 |
|  | 31 | 9.38 | 17.03 | 14.24 |
| 02-004 | 15 | 62.39 | 17.116 | 7.36 |
|  | -1 | 15.58 | 11.865 | 5.78 |
| 02-007 | -4 | 8.92 | 5.39 | 12.53 |
|  | 13 | 38.72 | 7.00 | 8.51 |
| 02-018 | 21 | 185.06 | 43.58 | 11.55 |
|  | -4 | 19.45 | 20.33 | 14.54 |
| 02-022 | 21 | 96.47 | 27.39 | 14.12 |
|  | -2 | 49.99 | 28.78 | 14.80 |
| 02-028 | 0 | 4.64 | 16.44 | 19.15 |
|  | 29 | 52.42 | 92.99 | 110.09 |
| 02-035 | -1 | 12.26 | 13.51 | 17.49 |
|  | 28 | 20.30 | 25.98 | 31.54 |
| 03-001 | -28 | 20.61 | 85.87 | 79.91 |
|  | 13 | 47.03 | 19.91 | 15.36 |
|  | 41 | 21.53 | 23.28 | 16.61 |
| 03-002 | -26 | 25.70 | 22.54 | 13.53 |
|  | 8 | 96.36 | 23.90 | 15.58 |
|  | 16 | 22.86 | 22.33 | 12.11 |
| 03-004 | -22 | 11.55 | 24.82 | 20.04 |
|  | 32 | 31.66 | 34.23 | 21.84 |
| 03-005 | -20 | 19.21 | 22.84 | 14.92 |
|  | 15 | 46.14 | 25.04 | 16.45 |
|  | 36 | 39.17 | 27.14 | 22.49 |
| 03-006 | -16 | 34.39 | 49.86 | 37.53 |
|  | 13 | 55.45 | 35.49 | 27.50 |
|  | 33 | 34.40 | 37.74 | 29.29 |
| 03-008 | -1 | 27.85 | 14.37 | 9.80 |
|  | 26 | 20.50 | 14.10 | 12.53 |
| 03-009 | -27 | 33.73 | 29.82 | 11.93 |
|  | 43 | 17.30 | 24.77 | 4.25 |
| 03-010 | -51 | 20.02 | 29.82 | 11.93 |
|  | 17 | 22.98 | 33.32 | 9.24 |
|  | 32 | 29.00 | 24.77 | 4.25 |
| 03-011 | -6 | 20.80 | 34.59 | 13.98 |
|  | 29 | 19.00 | 44.80 | 18.28 |
| 03-012 | -8 | 14.01 | 10.81 | 15.26 |
|  | 28 | 37.45 | 11.28 | 16.69 |
| 04-003 | -5 | 7.76 | 19.10 | 11.76 |
|  | 28 | 26.54 | 20.83 | 13.97 |
| 04-004 | 0 | 13.05 | 16.22 | 15.09 |
|  | 27 | 13.56 | 27.36 | 15.55 |
| 04-009 | -1 | 24.24 | 24.96 | 23.59 |
|  | 19 | 85.01 | 32.48 | 13.94 |
| 04-012 | -9 | 11.02 | 20.61 | 18.58 |
|  | 35 | 17.39 | 19.86 | 15.48 |
| 04-014 | -1 | 12.84 | 24.00 | 22.73 |
|  | 30 | 18.82 | 17.31 | 17.79 |
| 04-016 | -1 | 37.21 | 19.25 | 10.62 |
|  | 40 | 51.00 | 16.13 | 10.27 |
| 04-019 | -1 | 9.98 | 13.64 | 5.86 |
|  | 16 | 17.99 | 11.32 | 2.90 |
| 04-021 | 0 | 17.49 | 11.12 | 35.27 |
|  | 28 | 58.96 | 23.47 | 20.79 |
| 04-032 | -15 | 6.00 | 22.20 | 18.11 |
|  | 21 | 9.23 | 17.44 | 18.91 |
| 04-037 | -1 | 11.31 | 21.10 | 4.48 |
|  | 44 | 48.63 | 50.65 | 25.03 |

Suppl. Table 1: Cir-nDNA, MPO and NE values detected in samples of patients. Cir-nDNA: circulating nuclear DNA; MPO – myeloperoxidase, NE - neutrophils elastase, where 0 –date of surgery.

Supplementary Table 2

**A**

| Days | n patients | Median | Mean | SD | CV, % |
| --- | --- | --- | --- | --- | --- |
| 1-7 | 13 | 3,08 | 3,7 | 1,97 | 52,77% |
| 8-14 | 11 | 2,28 | 3,7 | 4,85 | 130,12% |
| 15-21 | 11 | 1,93 | 3,0 | 2,55 | 85,90% |
| 22-28 | 15 | 1,11 | 1,5 | 0,96 | 64,38% |
| 29-35 | 17 | 1,12 | 1,8 | 2,52 | 140,55% |
| 36-42 | 11 | 1,09 | 1,2 | 0,55 | 43,98% |
| 43-49 | 8 | 1,65 | 2,2 | 1,88 | 86,84% |

**B**

| Days | n patients | Median | Mean | SD | CV, % |
| --- | --- | --- | --- | --- | --- |
| 1-7 | 13 | 3,08 | 3,41 | 1,30 | 38,23% |
| 8-14 | 11 | 2,28 | 2,48 | 1,00 | 40,13% |
| 15-21 | 11 | 1,93 | 2,47 | 1,39 | 56,41% |
| 22-28 | 15 | 1,11 | 1,42 | 0,82 | 58,00% |
| 29-35 | 17 | 1,12 | 1,27 | 0,60 | 46,97% |
| 36-42 | 11 | 1,09 | 1,23 | 0,36 | 29,36% |
| 43-49 | 8 | 1,65 | 1,84 | 1,30 | 70,61% |

Suppl. Table 2: analysis the x-folding factor of circulating nuclear DNA for each week of follow-up: A – all values in series, B – excluding pairs of extreme low and high values. C – Overhaul cir-nDNA concentration medians before and after surgery over an 8-week period, Mann Whitney test.

SD – standard deviation, CV - coefficient of variation.

Supplementary Table 3

**A**

| Days post-surgery | Nb patients | Pre-surgery median cir-nDNA, ng/mL | Post-surgery median cir-nDNA, ng/mL | post > median HI, nb | post > median HI, % | post > pre, nb | post > pre, % | post > pre 2x fold, nb | post > pre 2x fold, % | post > pre 3x fold, nb | post > pre 3x fold, nb |
| --- | --- | --- | --- | --- | --- | --- | --- | --- | --- | --- | --- |
| 1-7 | 13 | 15.5 | 65.0 | 13 | 100% | 13 | 100% | 13 | 100% | 8 | 62% |
| 8-14 | 11 | 20.6 | 40.9 | 10 | 91% | 10 | 91% | 6 | 55% | 3 | 27% |
| 15-21 | 11 | 19.2 | 46.1 | 9 | 82% | 8 | 73% | 5 | 45% | 4 | 36% |
| 22-28 | 15 | 17.5 | 20.3 | 14 | 93% | 6 | 40% | 3 | 20% | 2 | 13% |
| 29-35 | 17 | 20.0 | 22.0 | 14 | 82% | 8 | 47% | 2 | 12% | 1 | 6% |
| 36-42 | 11 | 20.6 | 21.5 | 11 | 100% | 5 | 45% | 2 | 18% | 0 | 0% |
| 43-49 | 8 | 15.7 | 19.5 | 6 | 75% | 5 | 63% | 2 | 25% | 2 | 25% |
| 50-56 | 1 | 9.0 | 11.4 | 0 | 0% | 1 | 100% | 0 | 0% | 0 | 0% |

**B**

| Days post-surgery | Nb patients | Pre-surgery median MPO, ng/mL | Post-surgery median MPO, ng/mL | post > median HI, nb | post > median HI, % | post > pre, nb | post > pre, % | post > pre 2x fold, nb | post > pre 2x fold, % | post > pre 3x fold, nb | post > pre 3x fold, nb |
| --- | --- | --- | --- | --- | --- | --- | --- | --- | --- | --- | --- |
| 1-7 | 13 | 20.2 | 22.0 | 12 | 92% | 8 | 62% | 1 | 8% | 1 | 8% |
| 8-14 | 11 | 18.3 | 19.9 | 7 | 64% | 4 | 36% | 1 | 9% | 1 | 9% |
| 15-21 | 11 | 22.5 | 25.0 | 9 | 82% | 4 | 36% | 1 | 9% | 0 | 0% |
| 22-28 | 15 | 15.1 | 21.4 | 12 | 80% | 8 | 53% | 1 | 7% | 0 | 0% |
| 29-35 | 17 | 24.0 | 24.8 | 14 | 82% | 6 | 35% | 1 | 6% | 1 | 6% |
| 36-42 | 11 | 19.3 | 16.8 | 9 | 82% | 4 | 36% | 0 | 0% | 0 | 0% |
| 43-49 | 8 | 18.4 | 24.2 | 7 | 88% | 5 | 63% | 2 | 25% | 1 | 13% |
| 50-56 | 1 | 18.3 | 16.2 | 1 | 100% | 0 | 0% | 0 | 0% | 0 | 0% |

**C**

| Days post-surgery | Nb patients | Pre-surgery median NE, ng/mL | Post-surgery median NE, ng/mL | post > median HI, nb | post > median HI, % | post > pre, nb | post > pre, % | 2x fold, nb | post > pre 2x fold, % | post > pre 3x fold, nb | post > pre 3x fold, nb |
| --- | --- | --- | --- | --- | --- | --- | --- | --- | --- | --- | --- |
| 1-7 | 13 | 17.2 | 19.8 | 13 | 100% | 6 | 46% | 1 | 8% | 1 | 8% |
| 8-14 | 11 | 13.5 | 15.6 | 9 | 82% | 4 | 36% | 1 | 9% | 0 | 0% |
| 15-21 | 11 | 14.5 | 12.1 | 8 | 73% | 2 | 18% | 0 | 0% | 0 | 0% |
| 22-28 | 15 | 16.2 | 15.6 | 14 | 93% | 5 | 33% | 0 | 0% | 0 | 0% |
| 29-35 | 17 | 17.2 | 17.4 | 15 | 88% | 6 | 35% | 1 | 6% | 1 | 6% |
| 36-42 | 11 | 15.1 | 17.1 | 10 | 91% | 3 | 27% | 0 | 0% | 0 | 0% |
| 43-49 | 8 | 14.9 | 14.2 | 8 | 100% | 3 | 38% | 1 | 13% | 1 | 13% |
| 50-56 | 1 | 12.4 | 16.9 | 1 | 100% | 1 | 100% | 0 | 0% | 0 | 0% |

Suppl. Table 3: Comparative characteristics of circulating nuclear DNA (A), myeloperoxidase (B) and neutrophils elastase (C) values for patient groups in different periods of follow-up. Cir-nDNA: circulating nuclear DNA; MPO – myeloperoxidase, NE - neutrophils elastase.

Supplementary Table 4

| ID | cir-nDNA, ng/mL | MPO, ng/mL | NE, ng/mL |
| --- | --- | --- | --- |
| EFS 1520 | 10.3 | 8.34 | 8.62 |
| EFS 4969 | 8.9 | 12.41 | 7.59 |
| EFS 262 | 13.3 | 14.72 | 8.01 |
| EFS 3841 | 13.2 | 13.54 | 5.65 |
| EFS 5900 | 22.1 | 12.02 | 3.48 |
| EFS -385 | 6.9 | 16.63 | 11.51 |
| EFS 6161 | 15.0 | 13.88 | 8.46 |
| EFS 4567 | 18.5 | 14.03 | 7.88 |
| EFS 1061 | 6.5 | 12.41 | 11.37 |
| EFS 6153 | 12.4 | 24.18 | 7.69 |
| EFS 7469 | 9.3 | 14.13 | 5.57 |
| EFS 1109 | 11.7 | 8.29 | 3.35 |
| EFS 5935 | 12.9 | 9.23 | 9.20 |
| EFS 7739 | 11.5 | 9.52 | 6.10 |
| EFS 5927 | 27.9 | 15.50 | 14.16 |
| EFS 1539 | 11.7 | 9.08 | 5.81 |
| EFS 1555 | 14.9 | 12.36 | 5.36 |
| EFS 6196 | 13.5 | 13.34 | 5.97 |
| EFS 2531 | 9.9 | 4.23 | 3.53 |
| EFS 9005 | 16.5 | 13.15 | 9.94 |
| EFS 6188 | 38.8 | 15.70 | 10.08 |
| EFS 0624 | 7.7 | 4.42 | 4.06 |

| *Median* | *12.64* | *12.78* | *7.64* |
| --- | --- | --- | --- |
| *SD* | *7.22* | *4.19* | *2.81* |
| *Mean* | 14.25 | 12.32 | 7.43 |
| *CV%* | *50.67* | *34.00* | *37.77* |

Suppl. table.4: Determination of cir-nDNA, MPO and NE healthy individual median concentration values and positivity thresholds

Supplementary Table 5

**A**

| **Test de Mann-Whitney** | DATA Peri-THRuST VS HI | | Pre VS Post-surgery | |
| --- | --- | --- | --- | --- |
|  | p-value | p-value summary | p-value | p-value summary |
| Pre-surgery associated with "Post-surgery [1-7] days" | 0,1509 | ns | < 0,0001 | **** |
| Post-surgery [1-7] days | < 0,0001 | **** |  |  |
| Pre-surgery associated with "Post-surgery [15-21] days" | 0,0807 | ns | 0,0008 | *** |
| Post-surgery [8-14] days | < 0,0001 | **** |  |  |
| Pre-surgery associated with "Post-surgery [15-21] days" | 0,2855 | ns | 0,0759 | ns |
| Post-surgery [15-21] days | 0,0042 | ** |  |  |
| Pre-surgery associated with "Post-surgery [22-28] days" | 0,0847 | ns | 0,2671 | ns |
| Post-surgery [22-28] days | 0,0023 | ** |  |  |
| Pre-surgery associated with "Post-surgery [29-35] days" | 0,0621 | ns | 0,6098 | ns |
| Post-surgery [29-35] days | 0,0023 | ** |  |  |
| Pre-surgery associated with "Post-surgery [36-42] days" | 0,003 | ** | 0,4779 | ns |
| Post-surgery [36-42] days | < 0,0001 | **** |  |  |
| Pre-surgery associated with "Post-surgery [43-49] days" | 0,4692 | ns | 0,5054 | ns |
| Post-surgery [43-49] days | 0,0491 | * |  |  |
| Pre-surgery associated with "Post-surgery [50-56] days" | N=1, pas de statistiques possibles | | | |
| Post-surgery [50-56] days |  |  |  |  |

**B**

| **Test de Mann-Whitney** | DATA Peri-THRuST VS HI | | Pre VS Post-surgery | |
| --- | --- | --- | --- | --- |
|  | p-value | p-value summary | p-value | p-value summary |
| Pre-surgery associated with "Post-surgery [1-7] days" | 0,006 | ** | 0,5446 | ns |
| Post-surgery [1-7] days | < 0,0001 | **** |  |  |
| Pre-surgery associated with "Post-surgery [15-21] days" | 0,0042 | ** | 0,8977 | ns |
| Post-surgery [8-14] days | 0,1409 | ns |  |  |
| Pre-surgery associated with "Post-surgery [15-21] days" | 0,0037 | ** | 0,519 | ns |
| Post-surgery [15-21] days | 0,0013 | ** |  |  |
| Pre-surgery associated with "Post-surgery [22-28] days" | 0,0102 | * | 0,1703 | ns |
| Post-surgery [22-28] days | 0,0004 | *** |  |  |
| Pre-surgery associated with "Post-surgery [29-35] days" | < 0,0001 | **** | 0,8119 | ns |
| Post-surgery [29-35] days | 0,0002 | *** |  |  |
| Pre-surgery associated with "Post-surgery [36-42] days" | 0,0063 | ** | 0,519 | ns |
| Post-surgery [36-42] days | 0,0104 | * |  |  |
| Pre-surgery associated with "Post-surgery [43-49] days" | 0,0264 | * | 0,2345 | ns |
| Post-surgery [43-49] days | 0,002 | ** |  |  |
| Pre-surgery associated with "Post-surgery [50-56] days" | N=1, pas de statistiques possibles | | | |
| Post-surgery [50-56] days |  |  |  |  |

Supplementary Table 5 - continued

**C**

| **Mann-Whitney test** | DATA Peri-THRuST VS HI | | Pre VS Post-surgery | |
| --- | --- | --- | --- | --- |
|  | p-value | p-value summary | p-value | p-value summary |
| Pre-surgery associated with "Post-surgery [1-7] days" | < 0,0001 | **** | 0,9197 | ns |
| Post-surgery [1-7] days | < 0,0001 | **** |  |  |
| Pre-surgery associated with "Post-surgery [15-21] days" | < 0,0001 | **** | 0,6994 | ns |
| Post-surgery [8-14] days | 0,0011 | ** |  |  |
| Pre-surgery associated with "Post-surgery [15-21] days" | 0,0007 | *** | 0,4779 | ns |
| Post-surgery [15-21] days | 0,0107 | * |  |  |
| Pre-surgery associated with "Post-surgery [22-28] days" | < 0,0001 | **** | 0,8063 | ns |
| Post-surgery [22-28] days | < 0,0001 | **** |  |  |
| Pre-surgery associated with "Post-surgery [29-35] days" | < 0,0001 | **** | 0,8384 | ns |
| Post-surgery [29-35] days | < 0,0001 | **** |  |  |
| Pre-surgery associated with "Post-surgery [36-42] days" | < 0,0001 | **** | 0,6994 | ns |
| Post-surgery [36-42] days | < 0,0001 | **** |  |  |
| Pre-surgery associated with "Post-surgery [43-49] days" | 0,0017 | ** | 0,9591 | ns |
| Post-surgery [43-49] days | 0,0025 | ** |  |  |
| Pre-surgery associated with "Post-surgery [50-56] days" | N=1, no statistics available | | | |
| Post-surgery [50-56] days |  |  |  |  |

Suppl. Table 3: Statistical comparison of circulating nuclear DNA (A), myeloperoxidase (B) and neutrophils elastase (C) values before and after surgery for patient up to 8 weeks of follow-up. Mann Whitney test. Cir-nDNA: circulating nuclear DNA; MPO – myeloperoxidase, NE - neutrophils elastase.
