## Supplementary materials for "Post-surgery level of circulating DNA in stage III colon cancer patients: impact on the reliability of minimal residual disease detection"

**Supplementary information 1**

**Detailled analysis of Figure 2 A-H**

During the first week post-surgery (Figure 2A), all 13 patients displayed at least a twofold increase in cirDNA levels, as compared to their matched pre-surgery value. In the subsequent [8-14] days post-surgery period (Figure 2B), 91% of the plasma samples showed elevated cirDNA levels. Among these, 27% experienced a twofold increase, while 55% demonstrated a threefold post-surgery increase in cirDNA concentration (Fig 2B and Suppl. ). During the third week post-surgery ([15-21] days, Figure 2C), 73% of the samples displayed elevated cirDNA levels, as compared to matched pre-surgery samples. Among these, 18% and 46% exhibited a two- and three-fold increase in cirDNA concentrations. Among the samples analyzed during the [22-28] days post-surgery period (Figure 2D), 47% showed higher cirDNA levels post-surgery, with 20% showing a two-fold increase, 20% demonstrating a three-fold increase, and 27% maintaining stable cirDNA levels. Analysis from the fifth post-surgery week ([29-35] days) showed 47% of samples with an increase in post-operative cirDNA levels, with 24% maintaining stable cirDNA concentrations, and 24% and 12% showing a two- and a three-fold increase in cirDNA levels (Figure 2E). 45% of samples tested within the sixth post-operative week ([36-42] days, Figure 2F) showed elevated cirDNA levels, with 18% and x% showing a two- and three-fold increase in cirDNA concentrations. Within the [43-49] days post-surgery timeframe (post-surgery seventh week, Figure 2G), 62% of patients had an increase in cirDNA concentrations, and 25% and x% of the samples showed a two- and three-fold increase. Lastly, the only plasma tested during the eighth week post-surgery ([50-56] days post-surgery Figure 2H) showed an x-fold increase in cirDNA level,as compared to the preoperative baseline value.
